# Different intervention strategies toward live poultry markets against avian influenza A (H7N9) virus: model-based assessment

**DOI:** 10.1101/2020.06.17.20133405

**Authors:** Guanghu Zhu, Min Kang, Xueli Wei, Tian Tang, Tao Liu, Jianpeng Xiao, Tie Song, Wenjun Ma

## Abstract

**Background:** Different interventions targeting live poultry markets (LPMs) have been applied in China for controlling the avian influenza A (H7N9), including LPM closure and “1110” policy (i.e., daily cleaning, weekly disinfection, monthly rest day, zero poultry stock overnight). However, the effects of these interventions have not been comprehensively assessed.

**Methods:** Based on the available data (including reported cases, domestic poultry volume, and climate) collected in Guangdong Province between October 2013 and June 2017, we developed a new compartmental model that enabled us to infer H7N9 transmission dynamics. The proposed model incorporated the intrinsic interplay among humans and poultry as well as the effects of absolute humidity and LPM intervention, in which different intervention strategies were parameterized and estimated by Markov chain Monte Carlo (MCMC) method.

**Results:** There were 258 confirmed human H7N9 cases in Guangdong Province during the study period. If without interventions, the number would reach 646 (95%CI, 575-718) cases. The temporal, seasonal and permanent closures of LPMs can substantially reduce transmission risk, which might respectively reduce human infections by 67.2% (95%CI, 64.3%-70.1%), 75.6% (95%CI, 73.8%-77.5%), 86.6% (95%CI, 85.7-87.6%) in total four epidemic seasons, and 81.9% CI(95%, 78.7%-85.2%), 91.5% (95%CI, 89.9%-93.1%), 99.0% (95%CI, 98.7%-99.3%) in the last two epidemic seasons. Moreover, implementing the “1110” policy from 2014 to 2017 would reduce the cases by 34.1% (95%CI, 20.1%-48.0%), suggesting its limited role in preventing H7N9 transmission.

**Conclusions:** Our study quantified the effects of different interventions and execution time toward LPMs for controlling H7N9 transmission. The results highlighted the importance of closing LPMs during epidemic period, and supported permanent closure as a long-term plan.

**Author summary:** Five waves of human influenza A (H7N9) epidemics affected China during 2013 and 2017. Its continuous emergence poses a big threat to public health. Given the key role of live poultry markets (LPMs) in H7N9 transmission, different interventions in LPMs (including the “1110’’ policy and LPM closure) were widely employed to prevent human infection with H7N9. Providing scientific evidence of their long-term effects is very important for the disease control, which can help to maximize control benefits and to minimize economic loss. To achieve this, we established a new transmission model and parameterized the intervention strategies. By using the proposed model to investigate the recent H7N9 outbreak in Guangdong Province, we quantified the effects of temporal, seasonal and permanent PLM closures, and the “1110’’ policy, as well as different intervention timing on the emergence of human H7N9 infections. The results can offer useful information for local authorities to take proper management in LPMs, and help in preparing optimal control strategies.

## Introduction

A novel reassortant avian influenza A (H7N9) virus, characterized by severe pneumonia, acute respiratory distress syndrome, and high mortality rate [1], was first identified in east China in March 2013 [2]. Since then, human cases of H7N9 infection have periodically occurred in China, with a larger fifth epidemic wave during the winter and spring of 2016/17. As of June 2017, a total of 1533 human infections with 592 deaths have been reported to the WHO [3]. The continuous emergence of avian influenza A (H7N9) virus poses a big challenge to public health in China and around the world.

Epidemiological surveys showed that human H7N9 infections were associated with the exposure to live poultry or potentially contaminated environments, which was usually located in live poultry markets (LPMs) [4-10]. Further studies found that LPMs would support the maintenance, amplification and dissemination of avian influenza virus (AIV) [5-7, 11, 12], as different types of poultry and birds (such as chicken, ducks, pigeon, quail) are placed densely in LPMs and some of them are casually slaughtered there [13]. Under this circumstance, many environmental samples collected from LPMs were observed to be positive for H7N9 [6, 7, 11, 14], and strong positive association was found between H7N9 presence and local density of LPMs in China [9, 15]. Moreover, LPMs are very common in China. A survey conducted in Guangzhou in 2014 showed that 68% of families visited LPMs once a week even during the H7N9 epidemic period [14]. LPMs therefore played an important role in the H7N9 transmission in China [16-19].

Given the critical role of LPMs, interventions targeting them have become the most frequently adopted measures in response to H7N9 outbreak. The usually implemented interventions include routine intervention and LPM closure. Routine intervention emphasizes on cleaning and disinfecting LPMs. The representative one is the “1110” policy (i.e., daily cleaning, weekly disinfection, monthly rest day, zero poultry stock overnight), which has been carried out in Guangdong Province since 2015. According to the closing duration, LPM closures can be divided into three types: temporary, seasonal, and permanent closure. These strategies are widely employed in different regions. For example, LPMs in Yangtze River Delta were closed after the emergency of human H7N9 cases and were reopened when no case was reported [20, 21]. LPMs in Shanghai were closed from the dates of the Chinese New Year to April since 2014. These two operations are typical prototypes of temporary and seasonal closure. Permanent closure of LPMs is usually implemented in some developed regions, such as Singapore. To effectively and efficiently control the spread of H7N9 virus, an urgent and essential task is to quantify these interventions’ effects.

Through statistical models and epidemiological surveys, a few studies have investigated the potential association of LPMs closure with the reduction of AIV-isolation rates and H7N9-incidence in humans [16, 17, 20, 22-26]. Previous studies indicated that the adopted interventions (temporary closure) in different cities of China can reduce the risk of human infections with different efficiency [20, 21, 25, 26]. Other studies found that temporary closure and disinfection of LPMs could reduce H7N9 virus contamination [14, 17, 23, 27], but not completely halted the persistence and dissemination of the virus, and long-term benefits were unremarkable [17, 23, 24]. Existing studies had several limitations that are worthy of further discussion: (i) These studies mainly focused on temporary LPM closure, but the “1110’’ policy were seldom evaluated; (ii) The study period of previous studies usually took up several months and it failed to infer the long-term effects of LPM interventions; and (iii) Seasonal factors (absolute humidity [AH] has been shown a strong negative correlation with the survival and transmission of influenza viruses [28-30]), were seldom incorporated into the modeling approach, which would influence the objectivity of assessment.

To fill the knowledge gaps, we conducted an empirical study by focusing the H7N9 transmission in Guangdong Province, where H7N9 epidemics have periodically emerged in recent years. We established a new compartmental model to explore the transmission dynamics. The effects of the “1110” policy and LPMs closure were fitted by Markov chain Monte Carlo (MCMC) method. By performing numerical simulations from the proposed model with the parameterized interventions, we quantified their specific impacts on the epidemic evolution, and based on that we put forward some intervention strategies.

## Methods and materials

### Study area

Guangdong Province (20°13’N to 25°31’N and 109°39’E to 117°19’66 E) is located in southern China with the most population (over 107.24 million) in China. It has an area of 179,800 km^2^, which is administratively divided into 21 prefecture-level cities (see Fig 1). Guangdong has a humid subtropical climate, with short, mild, dry winters and long, hot, wet summers. People there have the habit of purchasing live poultry at LPMs. The province accounts for about 10% of domestic poultry (DP) industry and it was regarded as an important epicenter of AIV circulation in China [13, 24].

**Fig 1.**
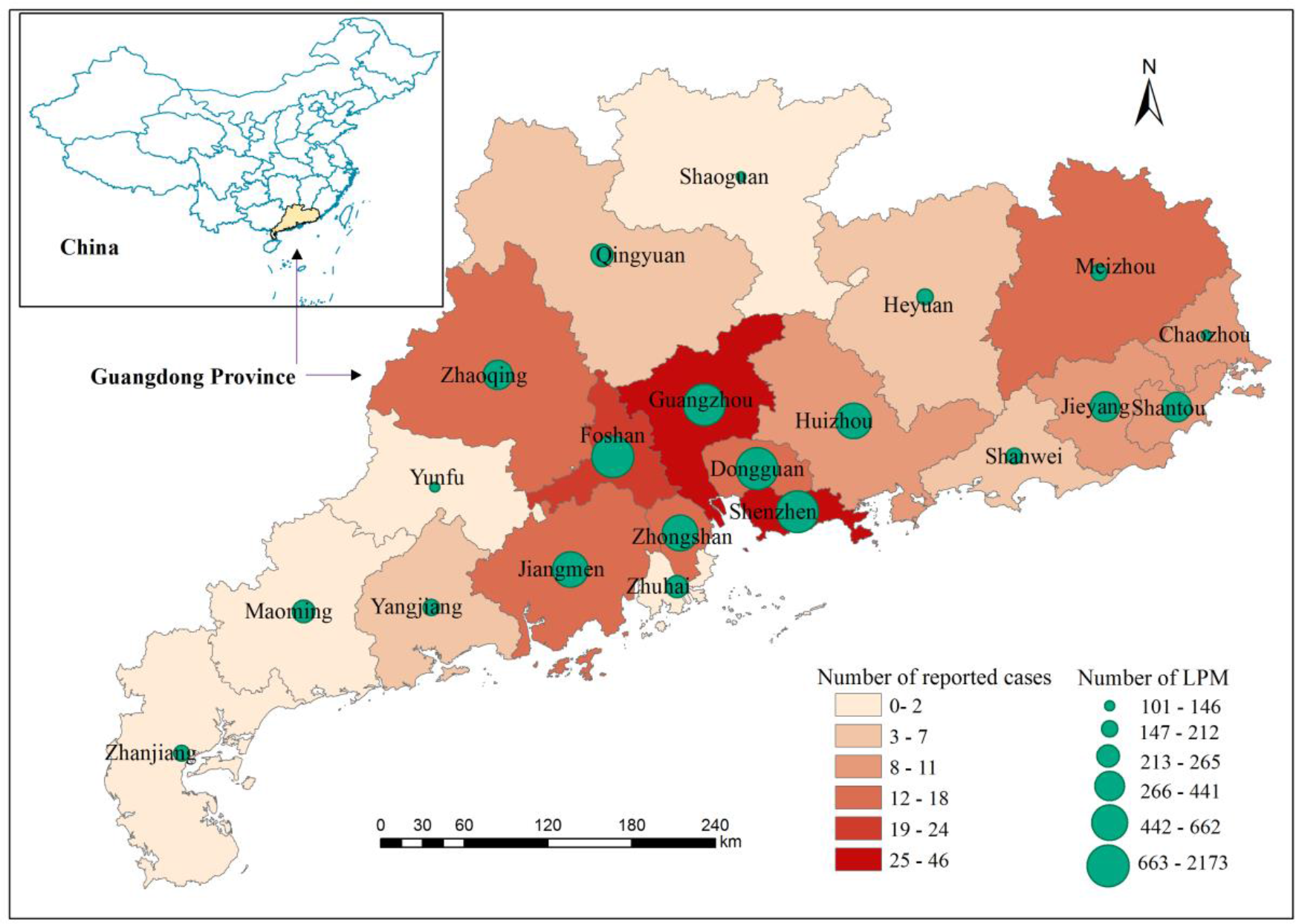
**The location of Guangdong Province in China, with the geographical distributions of reported H7N9 cases and live poultry markets (LPMs) in the 21 prefecture-level cities**.

### Data collection

The data of human H7N9 cases reported in Guangdong Province between October 2013 and June 2017 was used in this study. Records of weekly cases were extracted from Guangdong Provincial Center for Disease Control and Prevention (GDCDC), which include the date of illness onset and the city he/she lives.

The intervention strategies under consideration include the “1110” policy and LPM closure. The former was adopted by the Guangdong government and was enforced through this province since January 2015. To quantify the LPM closure data, we first retrieved the distributions of LPMs in city level by searching them from Baidu map, and then collected the numbers of closed LPMs as well as the closing time and duration by browsing local government webs and media reports (such information was documented by local government and delivered to the public by various media). Based on that, we computed the proportions of the closed LPMs in each week.

Weekly data of average temperature and relative humidity were retrieved from Guangdong Meteorological Service (http://www.grmc.gov.cn/), which were used to calculate the absolute humidity (AH). The seasonal numbers of the DP marketing were downloaded from the agricultural statistics provided by Statistics Bureau of Guangdong Province (http://www.gdstats.gov.cn/tjsj/ny/). The data was used to estimate the weekly recruitment rate of DP, i.e., the variable *A* in model (1). The original data of AH and DP is periodic and discontinuous. To specify these data to each week, the time series of AH and DP were smoothed by Fourier functions before applying them into the proposed model (see Figs S1 and S2).

### Dynamic model

Inspired by recently-developed mathematical models [31-35], we proposed a new compartmental model for simulating the H7N9 transmission dynamics among DP and humans, under the influence of LPMs closure and the “1110” policy as well as AH. Based on the compartmental principle and H7N9 epidemiology, we posed the following hypotheses.

- The total number of DP (N_p_) is classified into two subclasses: the susceptible (S_p_) and the infectious (I_p_); and the total number of humans (N_h_) is classified into four subclasses: the susceptible (S_h_), the exposed (E_h_), the infected (I_h_) and the recovered (R_h_).
- The transmission process follows an SI-SEIR framework. Thereinto, susceptible humans and poultry can be infected with certain probability once interacting with infectious poultry. If infected, human individual becomes ill after passing through the incubation period until recovery or death, and poultry becomes infectious immediately until death.
- The direct transmission among human beings is ignored, since there is no evidence for human-to-human infection [13, 33]. It is further assumed that the number of susceptible humans is static in each epidemic season (from October to next September) but varies across different seasons. The specific values were estimated by MCMC method.

Accordingly, the dynamical transmission flowchart is shown in Fig 2, and the corresponding model was formulated by the following ordinary differential equations.

**Fig 2.**
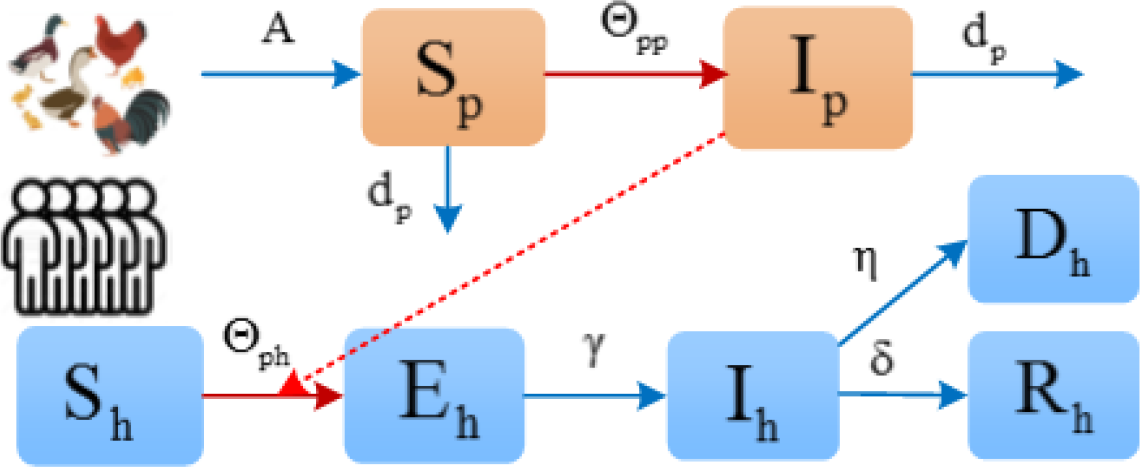
Flow diagram on the dynamical transmission of H7N9 infection among domestic poultry (DP) and humans. The transmission follows an SI-SEIR approach, where humans and poultry would be infected through direct/indirect interaction with infectious poultry.

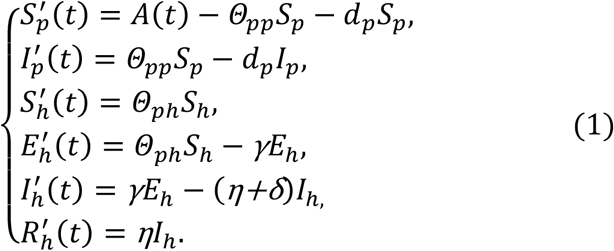

The interpretations of the variables and parameters are presented in Table 1. Here *Θ*_*pp*_ and *Θ*_*ph*_ represent the forces of infection from infectious DP to susceptible DP and to susceptible humans, respectively, given by:

**Table 1:**
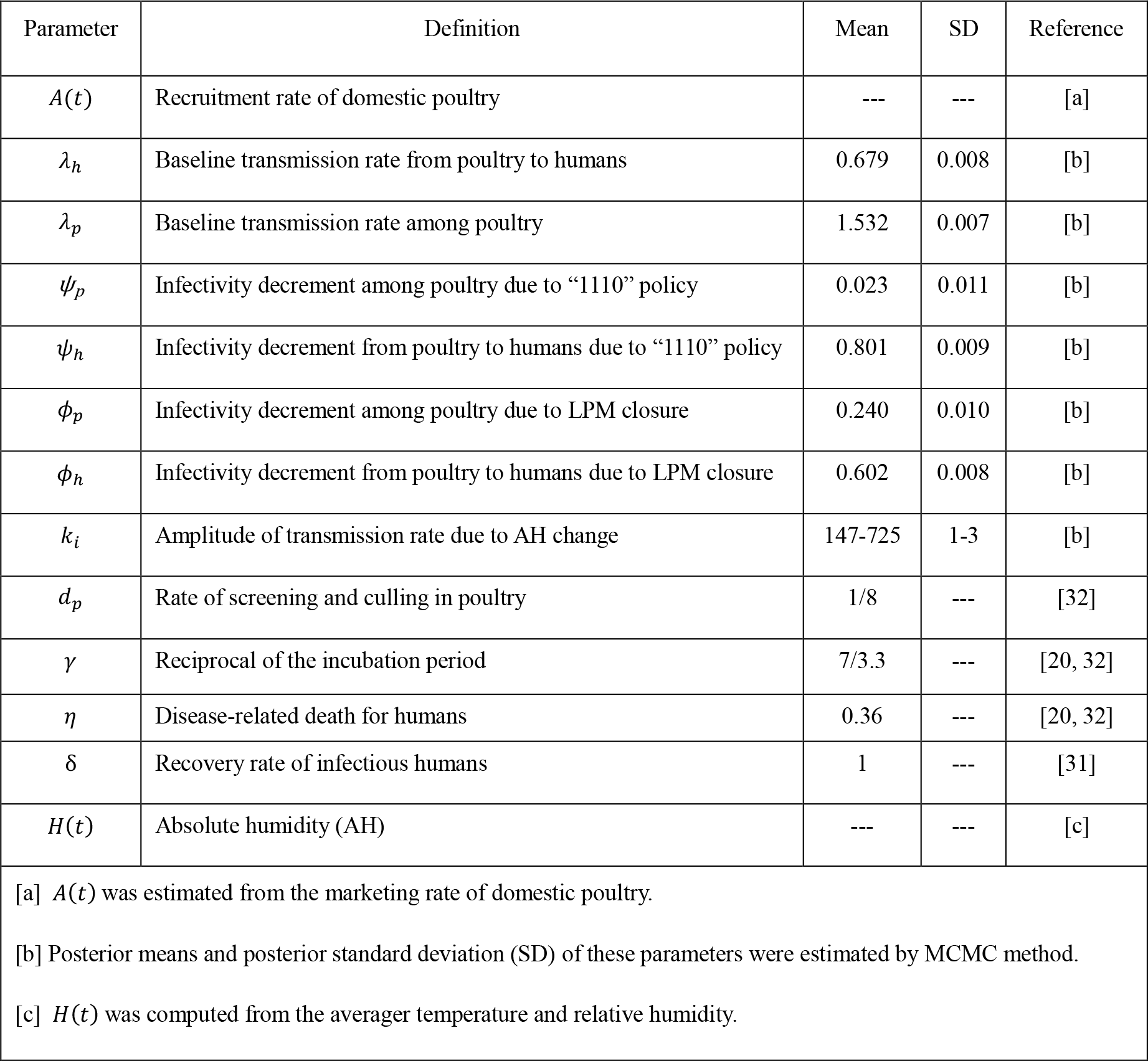
Description of parameters and variables in the model. The time unit is per week.

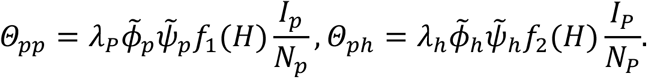

The above expressions include three components: (i) baseline transmission rates *λ*_*p*_ and *λ*_*h*_; (ii) effects of intervention strategies (i.e., LPMs closure and “1110” policy), measured by 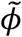 and 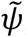; and (iii) seasonal influence, which is quantified by functions of AH, denoted by *f*_1_ and *f*_2_.

Since AH is negatively associated with H7N9 infections [28-30], to precisely estimate the role of AH in infectivity, we provided different expressions of *f*_1_ and *f*_2_ as follows.

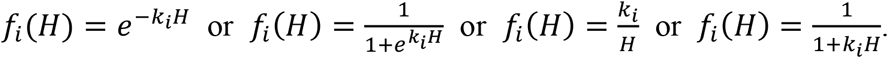

The selection of *f*_1_ and *f*_2_ is based on their performance in model simulations.

Since LPMs closure and the “1110” policy would reduce the contact rates (between humans and poultry, and within poultry) and infection probability [20, 24], the roles of these interventions were quantified by modulating the infectivity, In view of the fact that such strategies can weaken the interactions among poultry but could not cut off all of their connections (poultry would not stay alone), we proposed the following formulas:

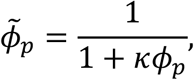

in case of κ proportion of LPMs being closed, and

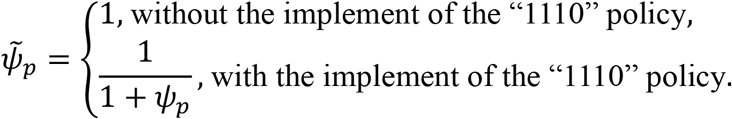

Furthermore, considering that these two strategies can reduce the interactions between humans and poultry more directly and significantly, we proposed the following formulas:

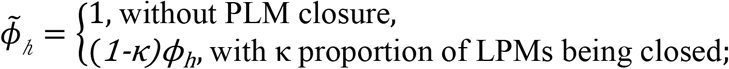

and

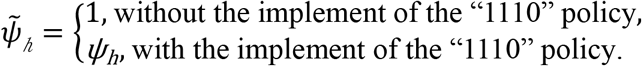

### Inferring model parameters

We adopted Metropolis-Hastings MCMC algorithm to estimate the parameters *Ω=*(*λ, ψ, Φ, k, S*_*h*_, *I*_*p*_(0)), by fitting the cumulative number of reported cases to modeling cases. Specifically, the relationship between reported case (I) and modeling cases (Y) was written as Y = I + ε, where Y=(Y_1_,Y_2_,…Y_T_) and I=(I_1_,I_2_,…I_T_) are

T-dimensinal vectors (T=196 is the study period). Each element in the random error vector ε follows normal distribution with zero mean and variance given by σ^2^. It is noted that Y can be generated by running the model (1) with given initial condition and parameters. Thus, the likelihood function of the estimated parameters can be written as:

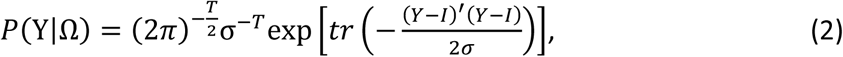

The corresponding joint posterior distribution of the estimated parameters is given by

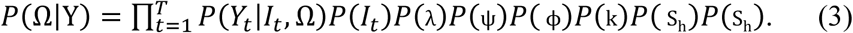

The procedure of the MCMC algorithm is handled as follows [36, 37]: First, all independent parameters Ω are initialized based on their prior information (following normal distributions). Second, the predictive cases are produced by solving the differential equations (1), and then the posterior distribution *P*(Ω|Y) is estimated by the equation (3). For each iteration, new values of parameters Ω^∗^ will be generated according to the adaptive proposal distribution *Q*(Ω^*∗*^|Ω), and will be accepted with probability

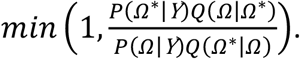

After a total of 20,000 iterations, the posterior distributions of the parameters are evaluated from the final 90% of the iterations.

To determine the most appropriate functional forms of *f*_1_ and *f*_2_, we substituted eight combinations of them into the model, and fitted each case by MCMC method. The criterion of model selection is measured by fitting errors (i.e., the mean square error [MSE]) and the Akaike information criterion [AIC]). Here

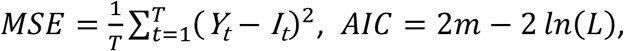

where *m* is the number of parameters and *L* is calculated by the equation (2).

### Uncertainty and sensitivity analyses

To assess the uncertainty in model output that roots in model input (i.e., parameters and initial conditions for the model variable), we used a Latin hypercube sampling (LHS) [38]. To further assess how variations in outputs can be apportioned to different input sources, we conducted sensitivity analysis by evaluating partial rank correlation coefficients (PRCCs) for different input parameters against output variables over time [38]. We performed these analyses by using LHS with 1000 samples. Because no data is available for the distribution functions of the input parameters, we chose normal distributions for them with mean values and standard deviations given by Table 1. The absolute value of PRCC clarifies the correlation between input parameters and output variables. The value greater than 0.4, between 0.2 and 0.4, or between 0 and 0.2, separately illustrates that the correlation is important, moderate, or not significant.

### Numerical analysis

For running the proposed model by ODE45 in Matlab, we presented the initial condition for the model as follows. The numbers of humans in exposed, infected and recovered states can be inferred from GDCDC, and the number of susceptible poultry was estimated from the marketing rate, which are E_h_(0)=I_h_(0)=R_h_(0)=0 and S_p_*=*2,146,293. While the initial values of susceptible humans S_h_(0) and infected poultry I_p_(0) were estimated by MCMC method.

To quantify the effects of different intervention strategies, we substituted the parameterized interventions into the model and performed a series of numerical simulations. The parameters vary according to the specific implementation of intervention at certain time. We first evaluated the influence of the adopted interventions, then checked the impacts of three types of closures as well as the “1110” policy, and finally assessed the intervention outcomes in case of different timing and duration. Here the duration of temporary, seasonal and permanent closure are assigned as from the following week of the first reported case to next January, from December to next February, from October 2013 to June 2017, respectively. The reduction rate ρ is calculated as *ρi*=1− *Θ*/*Φ*, where *θ* (*Φ*) is the number of human infections with (without) the implementation of intervention in the corresponding season.

## Results

A total of 258 human H7N9 cases with 100 deaths were reported in Guangdong Province between October 2013 and June 2017, in which four epidemic waves separately recorded 109, 72, 14 and 63 cases. The time series of the confirmed cases and the LPM intervention information are illustrated in Fig 3. It was observed that the epidemic curves exhibited seasonal cycles, in which human infections usually occurred in the winter/spring and peaked in January. In response to local infection, LPM closure was implemented in many areas of Guangdong Province for varying duration. It was found that parts of LPMs were closed from January, and the proportion of closed LPMs reached a maximum during the festival of Chinese New Year. In addition, the “1110” policy was carried out simultaneously in Guangdong Province from January 15, 2015.

**Fig 3.**
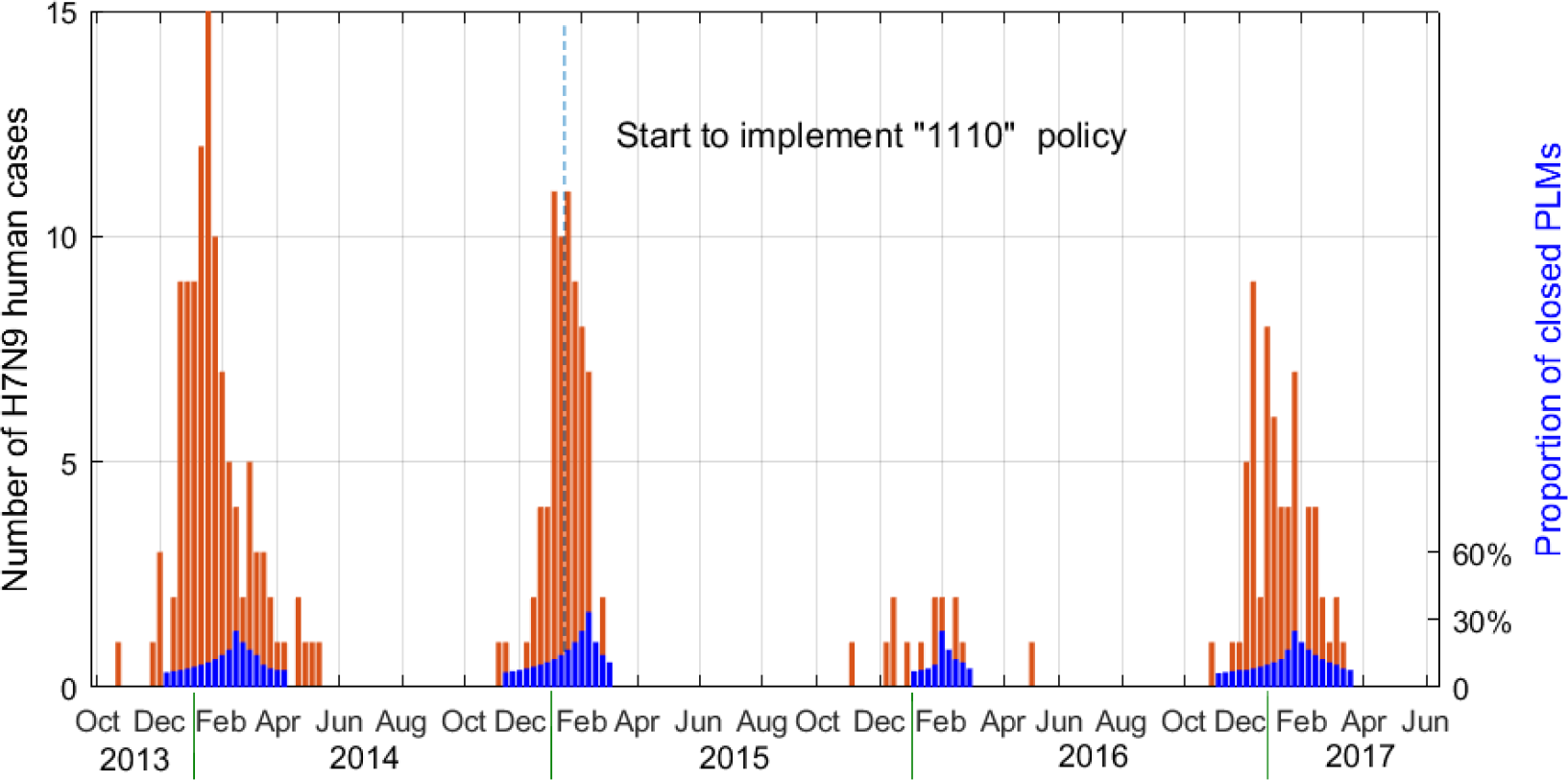
Weekly numbers of human infections with avian influenza A(H7N9) and the proportions of closed PLMs in Guangdong Province. Guangdong experienced four H7N9 epidemic waves from October 2013 to June 2017.

The time series of AH and DP volume also exhibited clear seasonal patterns (see Figs S1 and S2). As shown in Fig 4, regression analysis indicates that the onset of increased H7N9 incidence in Guangdong is associated with low AH levels and large DP volumes. Furthermore, the distributions of human infections in the 21 cities are in positive proportion to the numbers of LPMs, indicating the key role of LPMs in H7N9 transmission.

**Fig 4.**
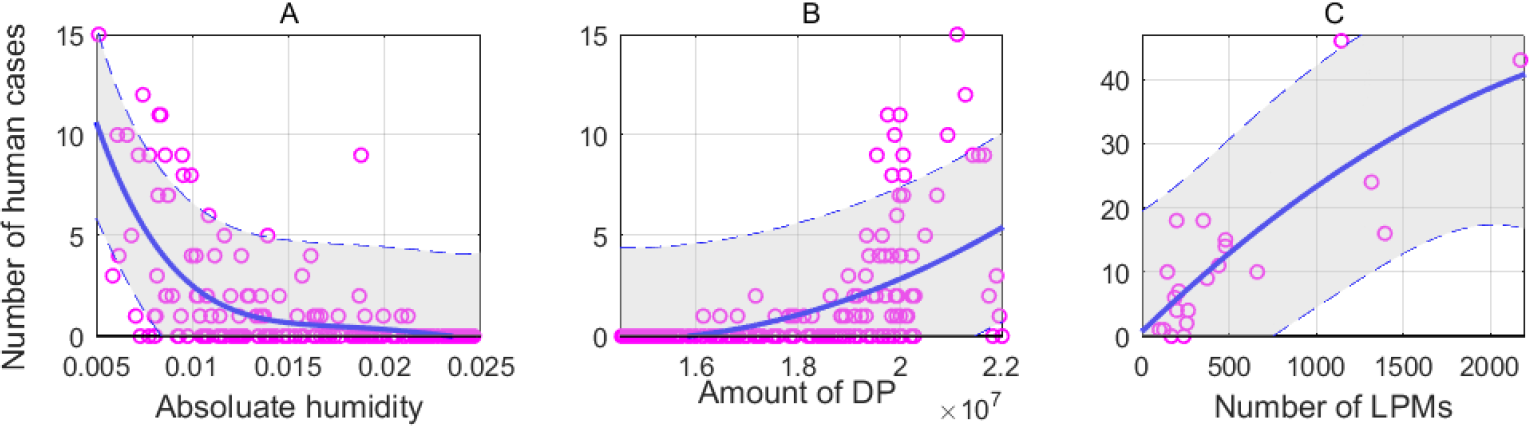
The association between the number of reported human H7N9 cases and absolute humidity, as well as the amounts of domestic poultry (DP) and LPMs. The data corresponds to the study period in Guangdong Province, with the unit as per week in (A) and (B) and as per city in (C). The light shaded area is the 95% confidence interval (CI).

We checked the performances of the model with different quantification of seasonal effects and then selected *f*_1_ and *f*_2_ as

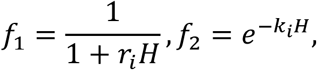

in that they yield the least values of MSE and AIC (see Table S1). In the following, we explored the outcomes of different interventions by applying the best-performance model to simulate H7N9 transmission dynamics.

The fitting results of weekly new cases and cumulative cases are shown in Fig S3. The proposed model performed well in fitting data, which accounts for 99.6% and 78.6% of variation in cumulative cases and weekly new cases, respectively (see Fig S4).

Fig 5 (A) shows the outcomes of the interventions that have already been adopted in Guangdong Province. The results indicate that during the study period if the “1110” policy had not been implemented in Guangdong, there would be 319 (95%CI, 286-351) cases of human H7N9 infection, and if the LPM closure had not been implemented, there would be 529 (95%CI, 457-602) cases. If both of these two kinds of interventions had not been implemented, the number would reach 646 (95%CI, 575-718) and the transmission would possibly continue until May in each year. In this case, the numbers of cases in the four epidemic curves could be 189 (95%CI, 183-195), 164 (95%CI, 151-176), 32 (95%CI, 27-36), and 265 (95%CI, 214-311), respectively. The simulation results show that the interventions adopted in Guangdong probably led to the reduction of human infection by 59.5% (95%CI, 48.5%-70.5%) and the prevention of 384 (95%CI, 313-456) cases.

**Fig 5.**
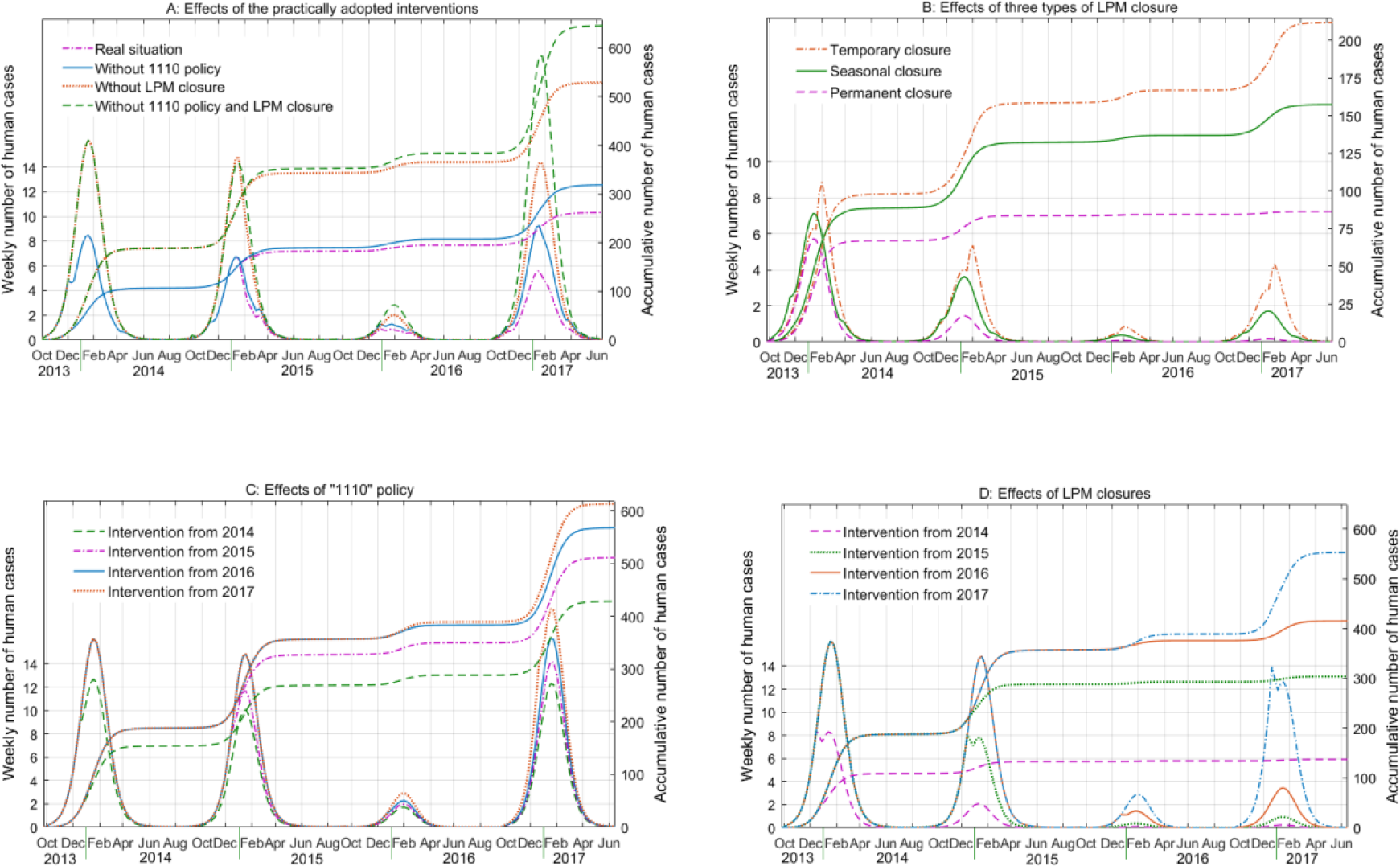
Estimated weekly incidence of human H7N9 infections with different intervention strategies, by employing the model to Guangdong Province. The simulation results correspond to (A) the practically adopted strategies; (B) the three types of LPM closure; (C) the “1110” policy and (D) LPM closure.

Figs 5 (B) and 6 display the impacts of the temporary, seasonal and permanent closures on human infection of H7N9. It is found that the three types of closure could dramatically reduce the magnitude of infections and shorten the epidemic period. Applying these strategies in Guangdong, numerical simulations show that the total number of human infections could be 212 (95%CI, 193-231), 157 (95%CI, 145-170) and 86 (95%CI, 80-93), which indicates 67.2%, 75.6% and 86.6% reductions in human infections, respectively. As shown in Fig 6, if temporary closure was implemented in the four epidemic seasons, there would be 48.0%, 63.2%, 74.3% and 83.0% decrease in human infections, compared to the situation without intervention in each season. In case of seasonal closure, the reduction rates would be 52.9%, 73.8%, 87.1%, 92.1%. These rates would be up to 64.6%, 90.2%, 96.8%, 99.2% for permanent closure, indicating its critical role in disease prevention. These data also indicate that if different LPM closure was implemented in four epidemic seasons, the control is more significant in the last seasons.

**Fig 6:**
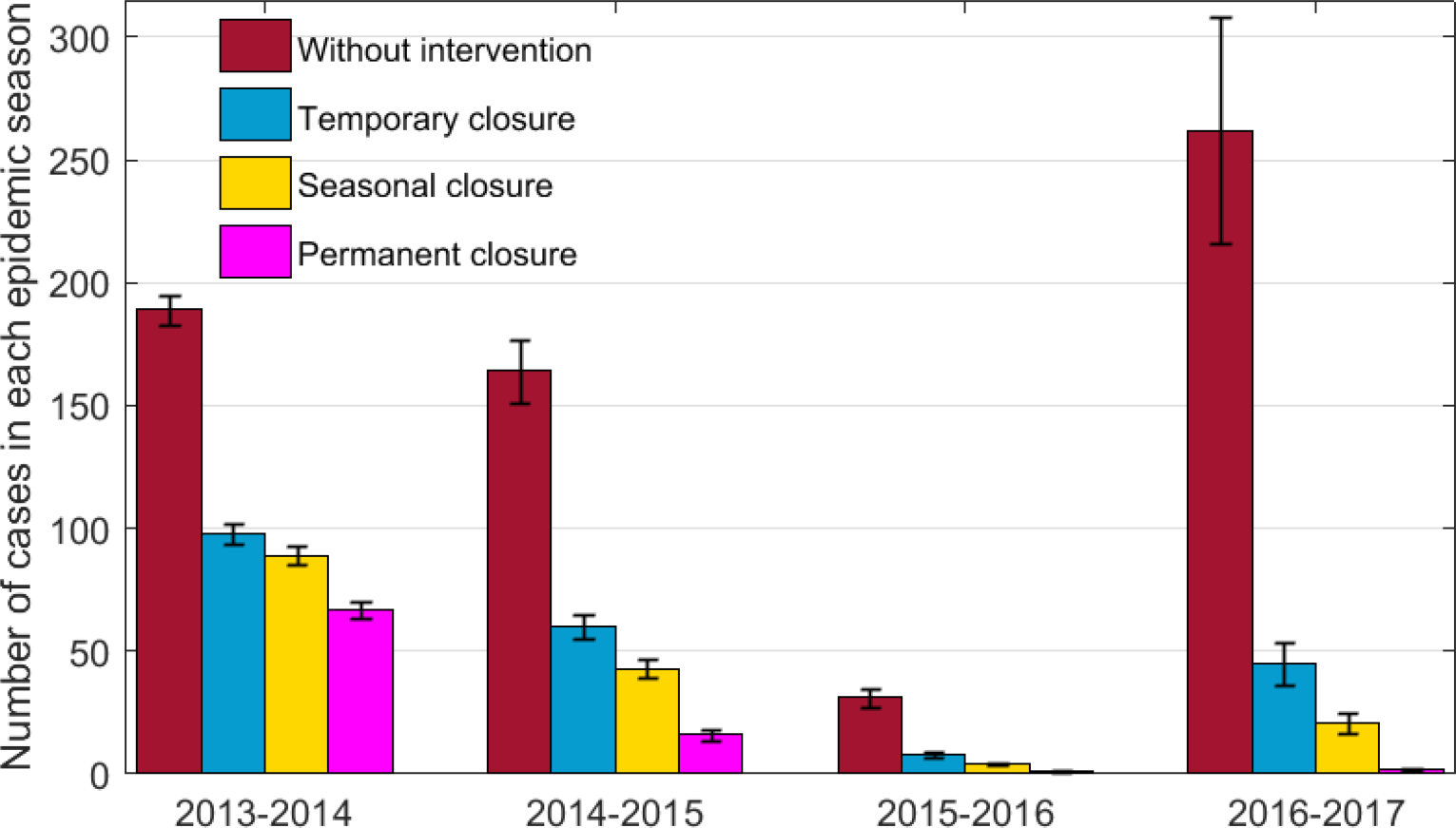
Estimated prevalence of human H7N9 infections with different types of LPM closure. The total numbers of human cases in each epidemic season with the 95% confidence intervals are shown.

Figs 5 (C and D) and Fig 7 show the comparison results of LPM closure and “1110” policy, as well as different start timings of interventions. It is found that these two interventions played a role in controlling H7N9 transmission, but closing LPMs was much more effective, which was associated with a sudden and sharp reduction in incidence. While the “1110” policy worked slowly and gently, and it could not eliminate infection risk in Guangdong. Moreover, the effects of interventions could be more pronounced if they were carried out earlier during the past five years. For example, if LPM closure or the “1110” policy were implemented since 2014, then 509 (95%CI, 502-516) or 220 (95%CI, 130-310) cases could be avoided. While if it was implemented since 2017, only 93 (95%CI, 44-142) or 33 (95%CI, 0-104) cases could be prevented.

**Fig 7:**
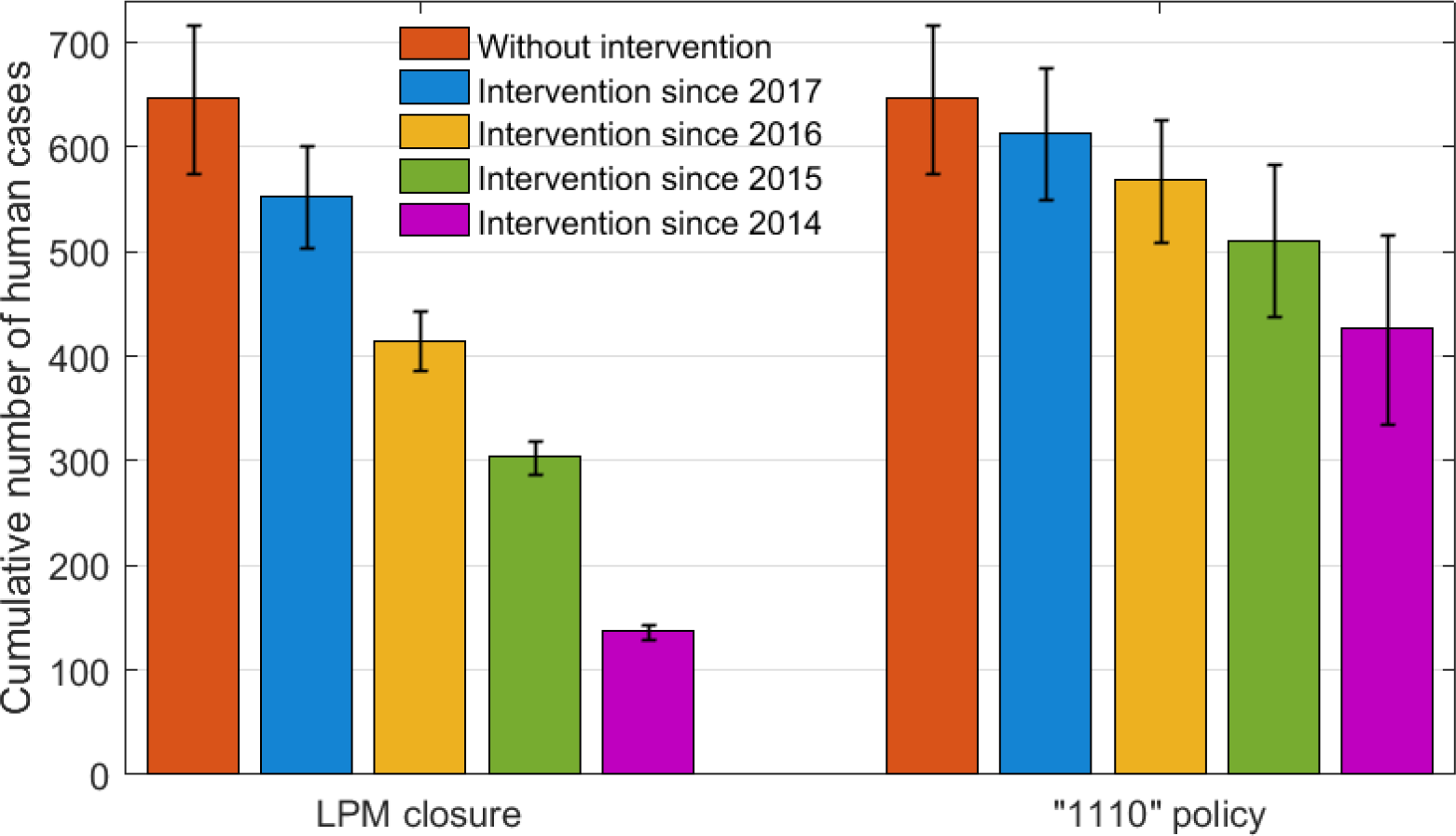
Estimated prevalence of human H7N9 infections with intervention beginning in different years. The cumulative numbers of human cases during four epidemic seasons with the 95% confidence intervals are shown.

Finally, we estimated the effects of interventions with different starting time and duration. Some patterns are observed in Fig 8: (i) Disease incidence keeps decreasing as prolonging the closure of LPMs, especially when the closure is between October and next February; (ii) Closing LPMs during the emergence of human infection always play a role in reducing the risk of infection, and the effects would be magnified if the “1110” policy is implemented in LPM-open days; (iii) For one month’s intervention, closing LPMs in every January receives the most benefits, which can reduce total infections by 41.1% (or 58.8%, with the combination of the “1110” policy); (iv) To most quickly achieve over 70% reduction in human infections, LPMs should be closed from early December to late February, and if combining the “1110” policy, this reduction rate could be above 75%; (v) Persistent intervention would bring much lower morbidity in latter epidemic seasons. For example, if LPMs were closed from November to next March in each year, there would be 79.5% (93.5%) reduction in human infections in the four (last two) epidemic seasons.

**Fig 8.**
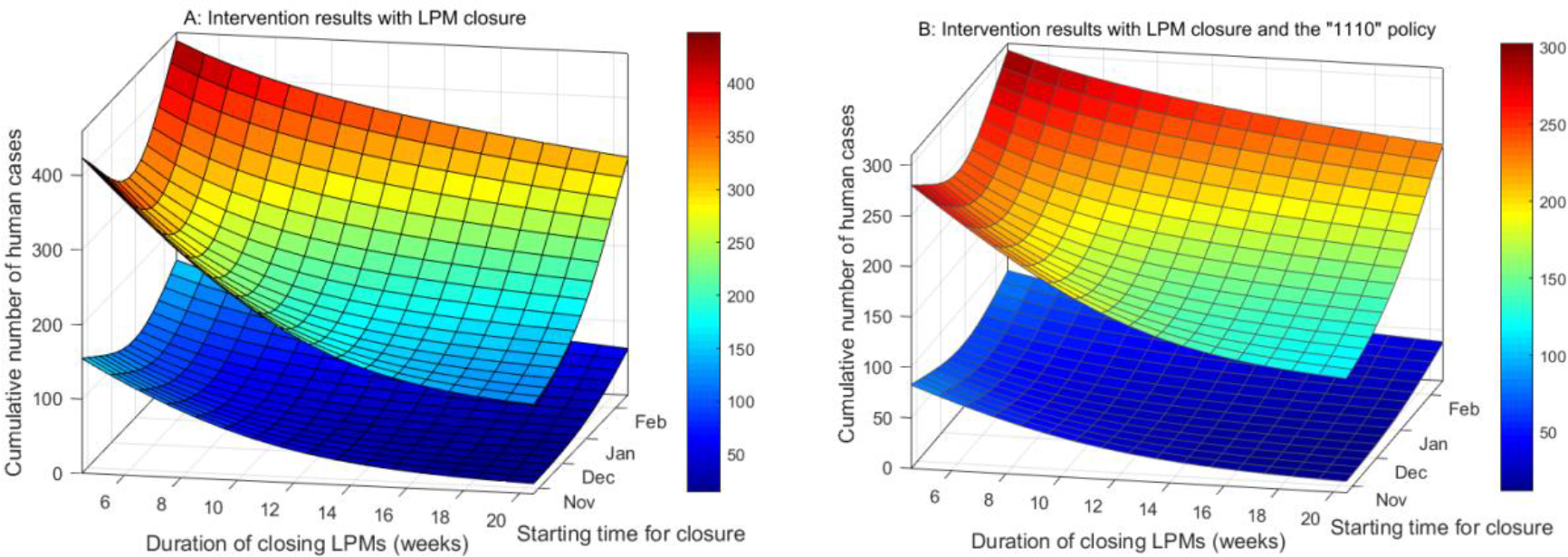
Estimated prevalence of human H7N9 infections with different intervention timing. The upper and lower curves are the numbers of human cases in the four epidemic seasons and in the last two epidemic seasons, respectively. The starting time of intervention varies from the last week of October to the second week of February. The duration of intervention varies from 5 weeks to 20 weeks.

Uncertainty and sensitivity analyses are shown in Figs 9, S5 and S6. Based on the absolute values of PRCC, the parameters that have strong influence in H7N9 transmission are the baseline transmission rates (*λ*_*p*_ and *λ*_*h*_), intervention indicators (*Φ*_*h*_, *Φ*_*p*_, *ψ*_*h*_ and *ψ*_p_), and seasonal index (*k*_1_ and *k*_2_). While the initial condition of the model is not sensitive in determining the model outputs (Fig S6). It is observed that the significance of these parameters varies over time: (1) The parameters with respect to the “1110” policy (*ψ*_*h*_ and *ψ*_*p*_) and LPMS closure (*Φ*_*h*_, *Φ*_*p*_) came into play only when they were implemented; (2) The baseline transmission rate from poultry to humans (*λ*_h_) and seasonal index become less and less correlated to the outputs; and (3) The baseline transmission rate among poultry (*λ*_p_) has consistent, major impact on the outputs.

**Fig 9.**
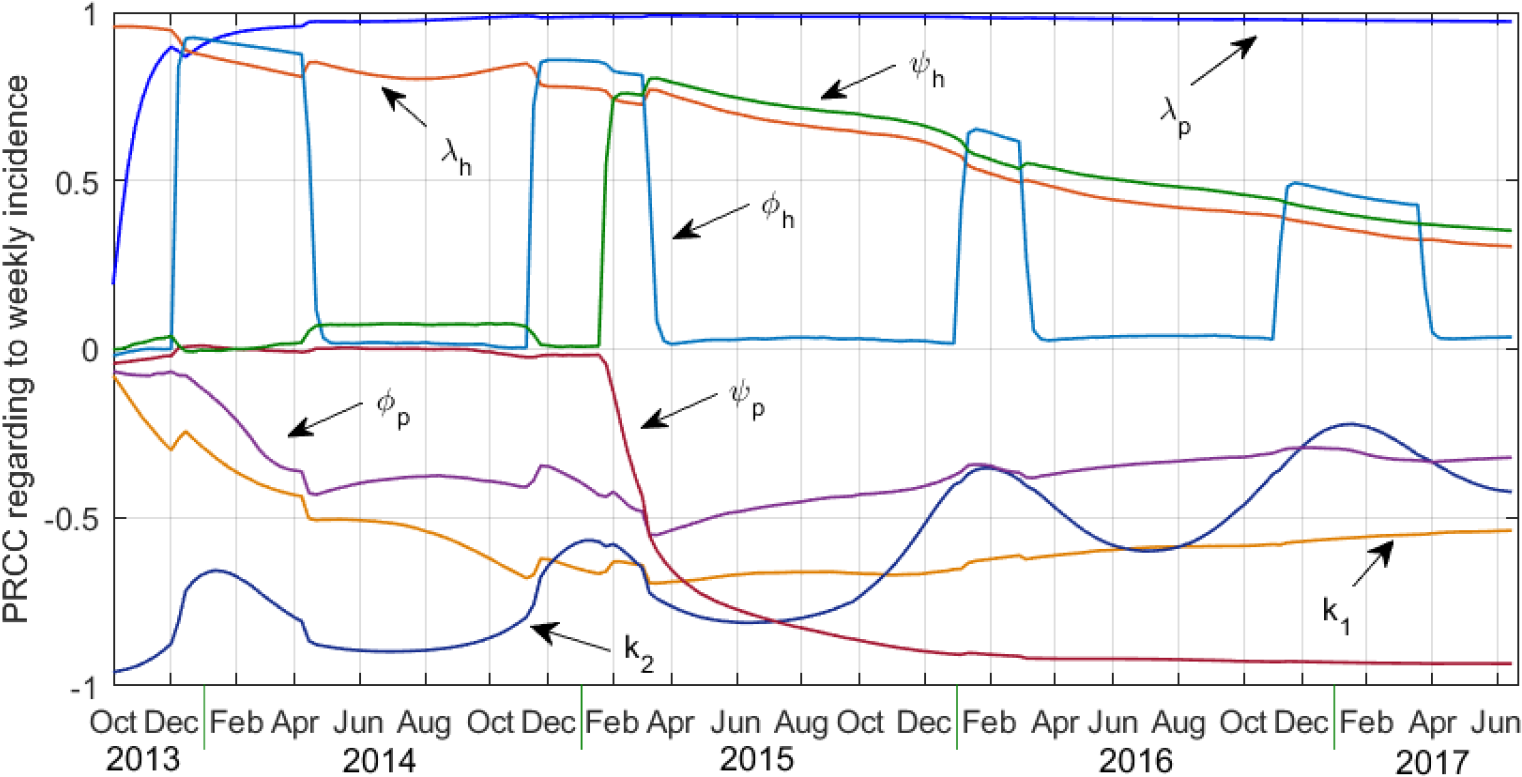
Time-varying sensitivity of the number of new human cases to the model parameters as indicated by PRCC values.

## Discussion

The present study established a new mathematical model to systematically assess the effectiveness of LPM management on human H7N9 infections, by applying the model to the recent outbreak in Guangdong Province. The proposed model incorporates the pivotal dynamic elements accounting for interactions between/among poultry and humans, and virus evolution in these two populations, as well as seasonality effect. These elements were intrinsically coupled by an SI-SEIR framework. Two important insights arising from our results are significant for policymakers to improve prevention strategies.

First, our findings indicate that the “1110” policy can reduce the intensity of H7N9 transmission, but it could not eliminate the risk of infection. Moreover, numerical analyses suggest that only implementing the “1110” policy is not enough to prevent human infection, but combining it with LPM closure (implementing them at different time) can perform much better for the control. The moderate effects of the “1110” policy based on our study is inconsistent with the studies conducted in Hong Kong [39, 40], where they observed substantial reduction in virus isolation rates and human cases after implementing a ban on overnight poultry storage and one rest day per month [39, 40]. The possible reasons could be the incomplete implementation of this policy in Guangdong, where the situation is more complicated and multifactorial. For example, in response to banning live poultry overnight, some sellers just moved or hidden the live poultry around the markets. In this regard, the real effects of the “1110” policy might be underestimated in our study.

Second, our findings predict that LPM closure can dramatically diminish human infections. It was estimated that the adopted interventions played a role in preventing human H7N9 infection in Guangdong Province. If the LPM closure was implemented uniformly in a larger area and persisted for longer period, the control effects would be much more significant. By comparing the incidence before and after LPM closures, several studies have found that closing LPMs could reduce the incidence by 97-99% in Eastern China in 2013 [20], 97% in Guangdong and Zhejiang during 2013-14 [25], and 73-99% in Shanghai, Zhejiang, Jiangsu and Guangdong [26]. The present study went a step further to examine the long-term transmission dynamic and considered that the temporal, seasonal and permanent closures of LPMs can effectively reduce human infections. Moreover, our study suggest that seasonal closure is more effective than temporal closure in preventing H7N9 transmission, but permanent closure achieves the best effectiveness. Another important issue for intervention is execution time. Numerical analyses imply that closing LPMs during the occurrence of H7N9 infection can maximize the efficiency of control. For example, closing LPMs from October 2013 to June 2017 or from December to March each year could result in 86.6% or 76.8% reduction in human infections, and these two reduction rates would be 99.1% and 92.8% in the last two epidemic season. Numerical results showed that the intervention strategies performed better in latter epidemic seasons. The possible reason could be that (1) a large number of poultry had been infected before the occurrence of human infection in 2013, and it took time to wipe out the infection source; and (2) persistent intervention plays a more significant role in reducing the infection risk.

Our results highlight the importance of LPM closure in epidemic seasons, and support the decision of closing LPMs in risky areas for preventing further human infections. The findings are consistent with the observations in Shanghai and Singapore, where there has been no zoonotic avian in humans after the respective implementation of seasonal and permanent closures of LPMs, and also verify the result found in [21] that reopening LPMs in summer may not increase the risk of human infection with H7N9.

At present China, due to the traditional poultry production-supply-consumption chain, temporally closing LPMs is at cost of huge economic loss. A recent study estimated that the temporal LPM closures in Shanghai, Hangzhou, Huzhou and Nanjing in 2013 cost the poultry industry about US$ 8 billion [10]. To minimize economic loss and to reduce the risk of infection, uniformly implementing seasonal LPM closure conducted by local government seems to be a sustainable intervention strategy, especially closing LPMs in the epidemic period combined with routine intervention in non-epidemic period. Furthermore, permanent closure should be considered as a long-term plan in China, since it is the most effective strategy to control H7N9 infection based on our study. Implementing this strategy needs to modify/improve the current poultry trade and live poultry market system in China, such as developing poultry farms instead of backyard poultry production, slaughtering and packaging poultry centrally in specialized factories, and selling frozen poultry instead of live poultry. These moves require combined efforts from the government and society, such as changing the cuisine culture away from preferring live poultry.

Several limitations exist in our study: (i) The proportion of closed LPMs in each week was estimated by the numbers of closed LPMs and the closing time, but the difference (e.g., scale, capacity) between these LPMs was not taken into account. (ii) The interventions were quantified by model parameters, the impact of which may be underestimated, given that the intervention in LPMs may not be implemented completely and effectively; (iii) The proposed model took into account the dominated routes for AIV transmission (including poultry-to-poultry and poultry-to-human transmission), but it did not incorporate the role of birds, which could be a potential factor that contributes to the spatial diffusion of AIV [33, 41]; (iv) Our analysis was based on a deterministic model with the potential effects of seasonality, which can captures the internal transmission mechanism and general trend of H7N9 infection, but it did not include the influence of random factors, such as environmental change or individual differences, and the results may contain certain uncertainties due to the data availability and model technology; (v) The temporal dynamics of AIV transmission under the influence of intervention was explored by the proposed model, but the model was unable to take into account the spatial heterogeneity and clustering of human infection, given the small number of cases and their sparse distributions in Guangdong Province; (vi) The impacts of vaccination was not considered here, which has dramatically reduced human infections since the use of vaccination in the second half of 2017.

## Conclusion

We have quantified the effects of different interventions and execution time toward LPMs for the prevention of human H7N9 infections by analyzing the transmission dynamics with a new and robust mathematical model. Based on the model simulation, we believe that the closures of LPMs (especially permanent closure) are highly effective in preventing H7N9 virus infections in humans, and emphasize the importance of closing LPMs from December to March in Guangdong Province. We further reckon that only implementing “1110” policy is unlikely to stop H7N9 transmission, and it should be combined with LPM closure in LPM-open days. The obtained results may provide useful information for local authorities to take proper management for LPMs, and help in preparing an optimal control strategy.

## Data Availability

The database is available from the corresponding authors upon request.

## Acknowledgments

This work was jointly supported by the National Natural Science Foundation of China (11661026), and Science and Technology Program of Guangzhou (201804010383).

